# DOSE: An open-source, iOS watch-based tool for experience sampling

**DOI:** 10.1101/2025.10.12.25337810

**Authors:** Ian Kim, Sahiti Kunchay, Saeed Abdullah, David E. Conroy

**Affiliations:** Department of Psychology, Stanford University, Stanford, CA, USA; Department of Pediatrics, Stanford University, Stanford, CA, USA; Department of Psychiatry, Yale University, New Haven, CT, USA; College of Information Sciences and Technology, Penn State University, University Park, PA, USA; School of Kinesiology, University of Michigan, Ann Arbor, MI, USA

**Keywords:** digital medicine, digital health, mobile health data, ESM, wearable computing

## Abstract

Smartwatches facilitate low-burden rapid-access micro-interactions, making them ideal for Experience Sampling Methods (ESMs). Despite the Apple Watch being the most popular smartwatch in the U.S., it has yet to be utilized in ESM studies due to a lack of accessible frameworks that enable deployment without technical expertise. We developed DOSE, an open-source ESM framework tailored for the Apple Watch. It includes tools and documentation that allow researchers to configure surveys, build custom apps, deploy studies, and stream data to servers without programming skills. We evaluated the framework’s feasibility in a 28-day field study with 18 participants (mean age = 55.3 ± 9.2). Results showed reliable prompt delivery and high response rates (>80% overall), with median interaction times under 10 seconds. Participants demonstrated increasing efficiency in responses over time. These findings establish the DOSE framework as a practical, scalable solution for Apple Watch-based ESMs and a foundation for future smartwatch research.

## INTRODUCTION

The Experience Sampling Method (ESM) is commonly employed to study individuals in their natural environments. ESM studies gather responses from repeated assessments at moments over time, from minutes to days to months, with the intent of obtaining lived, day-to-day experience of individuals, while minimizing retrospective biases that plague single-timepoint, between-person assessments (Chen, 2006; Gabriel et al., 2019; Shiffman et al., 2008; Stone & Shiffman, 1994). The technology on which this research method to solicit respondents’ answers at random moments of the day is based has changed extensively, moving from paper diaries to pagers to personal digital assistants to connected mobile devices (Barrett & Barrett, 2001; Hektner et al., 2007). However, the basic features of the method have remained essentially the same since its inception in the 1970s and its future evolution will likely be driven by technological advances that support remote data collection, monitoring, and intervention. Wearable devices, such as smartwatches, are one of the latest technological advances for ecological momentary assessments and interventions; however, existing applications have concentrated on Android, Fitbit, and maker applications because of constraints in Apple iOS. In this paper, we provide the code for Apple Watch-based experience sampling that overcomes a limitation in the Apple SDK to permit extended monitoring periods.

Originally, ESM studies relied on traditional pen-and-paper methods, often supplemented with devices such as pagers for signaling participants to record their thoughts or feelings on paper. Personal digital assistants integrated the prompting and recording functions in a single device and eliminated the need to manually enter data from paper records. The advent of smartphones has provided a more efficient solution by integrating notification delivery, questionnaire display and response storage or transmission into a single mobile device that can be connected to other devices or information sources. Consequently, recent ESM studies frequently employ dedicated smartphone applications (apps) or cloud-based services that deliver prompts at random intervals throughout the day through various methods, including calls, text messages, or push notifications. Nevertheless, the use of smartphones presents certain limitations. For example, smartphone devices are not always attached to the body. Depending on how a smartphone is carried (e.g., on hands, in a bag or pocket), participants may fail to notice the prompts altogether. Participants may also forget to carry their smartphones at times. Separation of the user from the portable device can reduce access to their smartphone and result in missed prompts or delayed responses which can be problematic when the approach is underpinned by assumptions that experiences are sampled at random. Another limitation is that smartphones themselves can be a source of distractions due to their multiple functionalities, potentially compromising participants’ engagement and compliance with ESM study protocols.

To overcome such limitations associated with smartphones, researchers have turned their attention to wearable devices, particularly smartwatches, as a potential alternative for ESM. Smartwatches offer several advantages over smartphones for this type of work. First and foremost, smartwatches are worn on the wrist, increasing proximity and potentially reducing response latencies. Increased proximity can improve sampling coverage by reducing the likelihood of systematically missing responses due to the smartphone not being accessible (e.g., while exercising, when the smartphone is stored in a desk drawer or purse). Improved coverage will yield more valid estimates of the momentary experiences and their dynamics. Previous studies have demonstrated higher compliance rates when using smartwatches for ESM (65% on smartphone vs. 76% on smartwatch), indicating improved participant engagement and adherence to data collection protocols (Ashbrook, 2010; Hernandez et al., 2016; Intille et al., 2016; Ponnada et al., 2017; Rouzaud Laborde et al., 2021). Smartwatches also have fewer distractions compared to smartphones (i.e., receiving fewer notifications and providing less access to social media apps), allowing participants to maintain focus on ESM prompts and responses. An additional advantage that distinguishes smartwatches from smartphones on ESM is their capability to enable assessment of micro-interactions (Intille et al., 2016). Micro-interactions are defined as interactions in which a user retrieves a device and completes a single-item questionnaire on it, with each phase being completed in just a few seconds. The use of micro-interactions can significantly reduce participant burden and enable more intensive sampling over time (Intille et al., 2016). Additionally, smartwatches have attracted attention for their ability to readily link and align self-report data with markers from other sensors within the watch, such as HealthKit data. This integration allows for a comprehensive and holistic understanding of the user’s health and well-being.

More than half of adults in the US own a smartwatch (Nagappan et al., 2024; Shandhi et al., 2024). However, the adoption of smartwatches generally and the use of smartwatches for ESM specifically are still at an early stage. The limited studies using smartwatches for ESM have largely focused on evaluating the feasibility of the approach. The few existing smartwatch-based ESM apps are typically only available on the Android platform or custom hardware (Kheirkhahan et al., 2019; Kim et al., 2022; Manini et al., 2019; Rouzaud Laborde et al., 2021; C. Stone et al., 2025; Volsa et al., 2022). None are available for the Apple Watch despite Apple Watch’s dominant position in the smartwatch industry, boasting over 115 million active users and a 59% market share in the United States (Cybatt, 2021; Statista, 2024). This discrepancy has resulted in the systematic exclusion of a large segment of the population from smartwatch-based ESM research.

The primary reason for the Android platform’s dominance in ESM studies is that, as of February 2024, there are some key features that are harder to implement on iOS-based smartwatch ESM apps compared to Android-based smartwatch apps. First, iOS imposes restrictions on the number of scheduled notifications an app can have concurrently. Specifically, the system maintains a maximum of 64 notifications that are closest to their scheduled prompting time; any notifications exceeding this limit are discarded by the system and are not delivered to the user. Assuming a frequency of six daily surveys, ESM studies would exhaust 42 notification slots within a single week. As a result, the study’s duration would be restricted to approximately two weeks before reaching the maximum threshold of 64 notifications. In contrast, Android devices do not encounter the same constraint due to the more flexible notification system available on that platform.

Second, iOS does not provide a built-in automatic random date/time selection function specifically tailored for sending notifications in ESM studies. In contrast, Android provides various APIs and features, including *AlarmManager, JobScheduler* and *WorkManager*, that enable developers to generate random timestamps with preset time intervals within a desired range and set up an automatic prompt delivery as periodic tasks. Manually assigning random schedules for study participants within the iOS app code is not recommended due to privacy risks, limited adoptability (i.e., ability to change schedules dynamically during the study to accommodate changes in participant availability, preferences, or study requirements) and limited scalability (i.e., difficulty in managing a large number of participants). Releasing an iOS app with participant IDs to the App Store or TestFlight also requires additional steps due to data protection and ethical concerns, even if the IDs were randomly generated by a computer (Apple Inc., n.d.-a). Implementing a remote configuration system where the schedules are stored on a server or content management system can be a better alternative. However, this approach adds complexity to the app architecture, requiring extra networking code and error handing for retrieval and synchronization of schedule data. Moreover, its heavy reliance on the server can cause issues where participants fail to receive schedules due to server downtime or connectivity problems.

Lastly, the closed-source nature of iOS has limited the extent of active discussions and comprehensive documentation surrounding app development processes. In contrast to open-source platforms like Android, where developers actively collaborate, share code snippets, and exchange ideas, the iOS ecosystem has fewer opportunities for community-driven discussions and knowledge sharing. Developing a new app from the scratch requires a significant investment of time and cost for researchers. The development process involves stages such as conceptualization, design, coding, testing, and deployment, which can span several months or even longer. Financial investment is also often necessary, particularly when it comes to expenses related to hiring skilled developers and app testers. This can pose a significant challenge for projects operating with limited budgets. Exploring existing and well-tested ESM app platforms can help reduce development time and costs, allowing researchers to tailor the app to their specific needs while minimizing study deployment delays. Unfortunately, as of now, there is no publicly available tool or customizable platform specifically designed for iOS smartwatch-based ESM research app development.

### The DOSE App

We designed and programmed DOSE, an open-source iOS smartwatch template application, to address the aforementioned issues. A key innovation of the app is that it enables researchers to work around the limitation of 64 notifications on iOS by implementing a system that dynamically manages the notification schedule within the app. The DOSE template contains streamlined code that determines the number of notifications to send each day, the number of days into the future for which notifications should be scheduled, and the interval for automatic rescheduling of notifications. If three notifications are set to be sent per day and 20 days into the future are scheduled, the total scheduled notifications would be 60. If an interval of 5 days is set, the app will automatically reschedule the next 60 notifications every fifth day. A shorter interval is beneficial for participants traveling frequently across different time zones. This approach ensures a continuous flow of notifications for participants enrolled in ESM studies without reaching the 64-notification limit. These notifications can be delivered to participants’ Apple watch at evenly distributed random times, within pre-determined time windows. Researchers can also easily adjust the parameters within the code to increase or decrease the number of surveys to be delivered per day (equivalent to the number of time windows per day as only one notification is sent per window) or adjust the time intervals for each window by simply updating the corresponding values. This flexibility empowers researchers to easily customize the notification (experience sampling) schedule according to their specific study requirements.

The DOSE framework was created using SwiftUI in Xcode. The back end of the current version of DOSE framework is based on Google Firebase Realtime Database as a data storage and a processing unit. With this framework, researchers have the flexibility to integrate various types of sensor data collection alongside ESM survey response data collection. As an illustrative example, the framework includes automatic collection of step counts and heart rate metric data through HealthKit during the predetermined study period. The full open-source code and conditions of use for this framework application are available for download via GitHub (https://github.com/iansulin/umich_dose). The workflow process for conducting an iOS smartwatch-based ESM study using the DOSE framework can be divided into three main stages: (1) compilation of the app to meet research design specifications, (2) participant onboarding and data collection, and (3) data processing for analysis.

### Step 1: App Customization

#### 1.1. Working Template

Following the installation of the necessary software (Xcode and Simulators; see https://developer.apple.com/documentation/safari-developer-tools/installing-xcode-and-simulators), a new project using this framework can be initiated by importing the project folder of the source code files into Xcode. This will create a basic working template application that can be customized for specific research purposes. The template includes a collection of tools: (1) `Utilities`, (2) `SurveyModules`, and (3) `Views`. Detailed explanations for functions and parameters can be found in the comments throughout the code.

#### 1.2. Utilities

The `Utilities` directory includes a range of essential components for the app, such as `survey`, `storage`, `notification`, `HealthKit`, and `Firebase manager systems`. Two crucial components that must be configured by individuals looking to utilize and integrate this framework app into a new, customized ESM app are the survey scheduling parameter definition and the database connection.

##### Database Configuration

The DOSE framework leverages the Firebase REST API to interact with the Firebase Realtime Database. By adopting the Firebase REST API, the framework eliminates the need for the `GoogleService-Info.plist` file and SDK configuration, which can be a daunting task for researchers without software development and programming expertise. When using the Firebase REST API, the only requirement is the Firebase Realtime Database URL. The URL typically follows the format “https://your-project-id.firebaseio.com” and can be found at the top of the Realtime Database section in the Firebase console (**Figure 1-a**). Within the `FirebaseManager`, there are three specific functions where the URL needs to be configured: uploading notification details, uploading HealthKit data, and uploading survey response data. These instances are marked with a placeholder text “`Replace with your Firebase Realtime Database URL`” (**Figure 1-b**).

**Figure 1.**
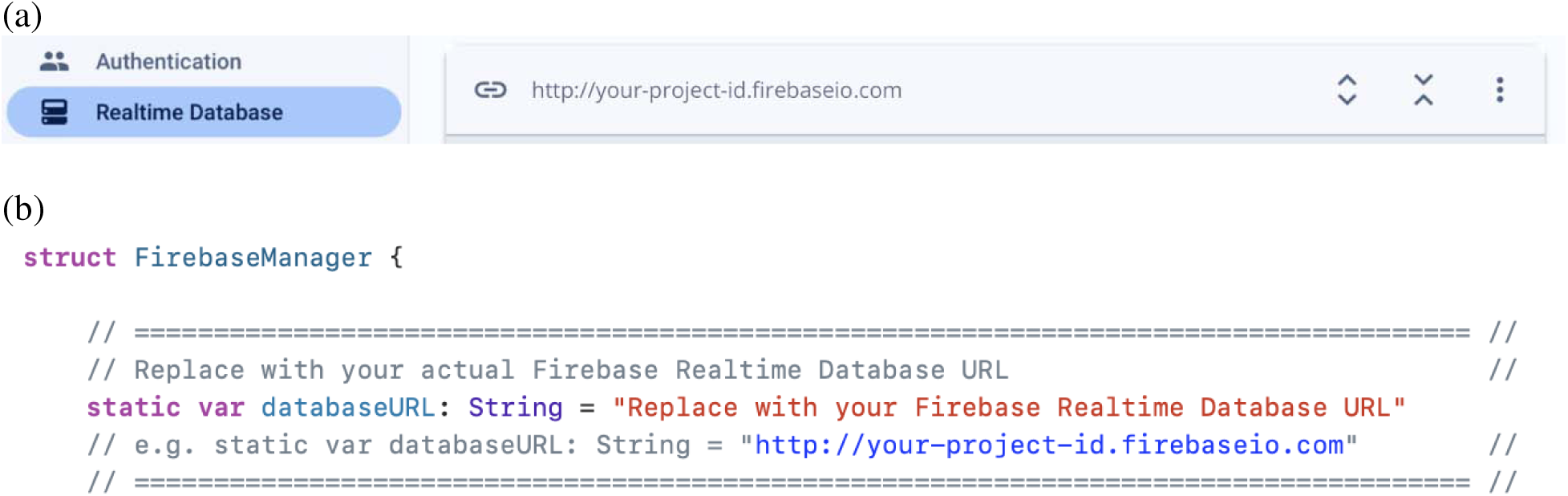
Database configuration

##### Survey Scheduling

Survey scheduling requires the specification of five numeric constant parameters in the `NotificationManager` file: `maxDayToSchedule`, `firstWindowStartHour`, `lastWindowEndHour`, `numberOfWindows` and `windowLength` (**Figure 2**). The `maxDayToSchedule` parameter determines the entire study period during which participants will receive survey notifications. It represents the number of days plus one, as survey prompt delivery begins on the day following participants onboarding. For example, if the study period is 180 days, the value of this parameter should be set to 181. The `firstWindowStartHour` and the `lastWindowEndHour` parameters define the earliest and latest times for survey prompt delivery. The specified time should be in a 24-hour format, without AM/PM. For instance, if prompts are to be delivered between 8 AM and 8PM, the parameters should be set as 8 and 20, respectively, rather than 8 and 8. The `numberOfWindows` and the `windowLength` parameters determine the number of windows per day and the length of each window. Windows are currently constrained to be a constant size across the day. The product of these two parameters should correspond to the duration between the earliest and latest times for survey prompt delivery. For example, if the earliest time is 8 and the latest time is 20, resulting in a 12-hour duration, and the length of each window is 4 hours, then the number of windows should be 3 to achieve a total duration of 12 hours. Depending on the specific requirements and features of the app, additional parameters may be added to customize the scheduling system.

**Figure 2.**
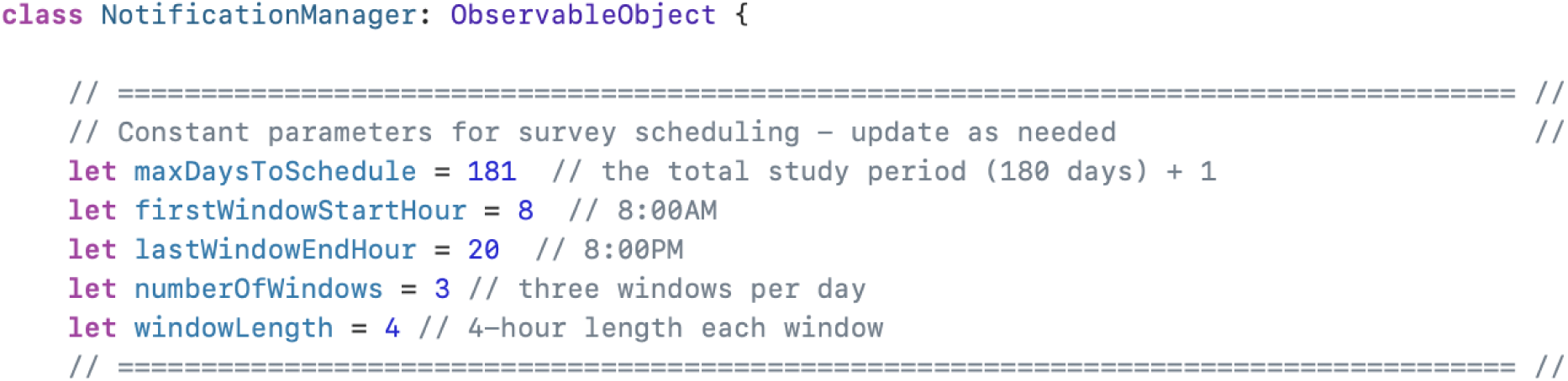
Survey scheduling.

#### 1.3. Survey Modules

##### Survey Interface

The `SurveyModule` directory comprises five survey interfaces (`rand_survey_1-5`), each featuring a distinct design with different layouts. Among them, two interfaces facilitate open-ended responses using user input. One interface incorporates a digit pad for numeric responses (`rand_survey_1`; **Figure 3-a**), while the other provides users with choices such as the regular Apple Watch keypad, scribbles, or voice typing for character or numeric input (`rand_survey_2`; **Figure 3-b**). Additionally, two interfaces offer slider scales. One employs a stepper slider, which allows individuals to increment or decrement a value using a two-segment control (`rand_survey_3`; **Figure 3-c**). The other employs a circular slider (`rand_survey_4`; **Figure 3-d**). The slider scale interfaces can be controlled through hand gestures (swiping or tapping) or by turning the Digital Crown, the rounded knurled knob on the side of the Apple Watch. Lastly, one interface contains a picker for multiple-choice questionnaires, that displays one or more scrollable lists of distinct values that users can choose from (`rand_survey_5`; **Figure 3-e**). Each survey interface includes two buttons at the bottom of the screen. The red button on the left enables users cancel the survey without submitting their response, while the green button on the right side of the screen allows users to submit their response.

**Figure 3.**
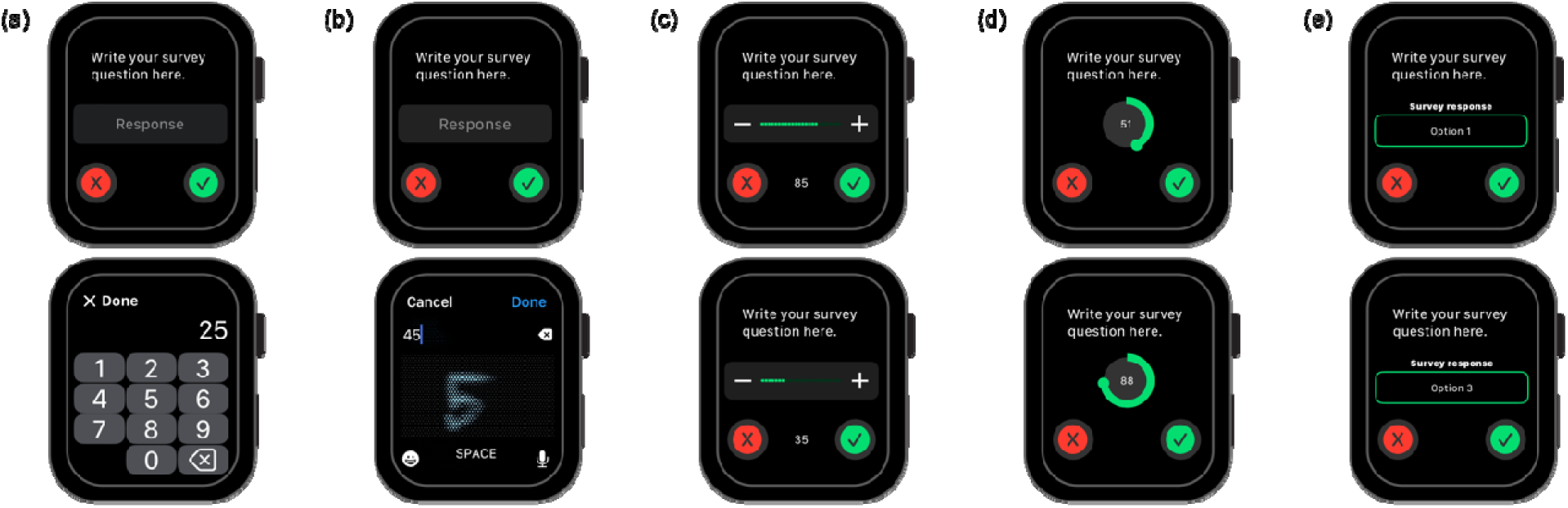
Input interfaces for the survey module: (a) Numeric Keypad, (b) Scribble/Dictation, (c) Step Slider, (d) Circular Slider, and (e) Scrollable List.

##### Survey Design

The survey question statements can be easily updated by replacing the placeholder text “`Write your survey question here`” under the commented line “// Survey question” in each survey module (**Figure 4-a**). For slider scales, the parameters for the minimum value, maximum value, and increment (for stepper slider) of the response can be modified under the commented line “`// Slider configuration`” (**Figure 4-b**). The picker interface lists can be updated by replacing the choice options under the commented line “`// Picker configuration`” (**Figure 4-c**). Each of the five survey modules is numbered from 1 to 5. When a user submits their responses, the survey response is sent to the survey database along with the corresponding survey module number, dates and times of survey delivery, opening, and submission. In cases where the survey was dismissed, a numeric value of `99999` is sent to the database as the survey response. If a participant opens a survey but submits it without entering any responses, the submission is still recorded, but the response field is left blank.

**Figure 4.**
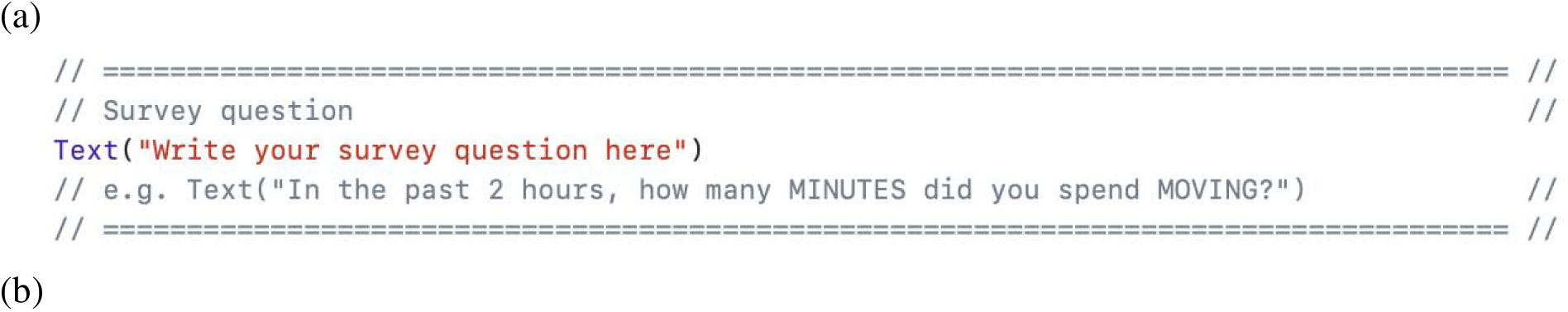
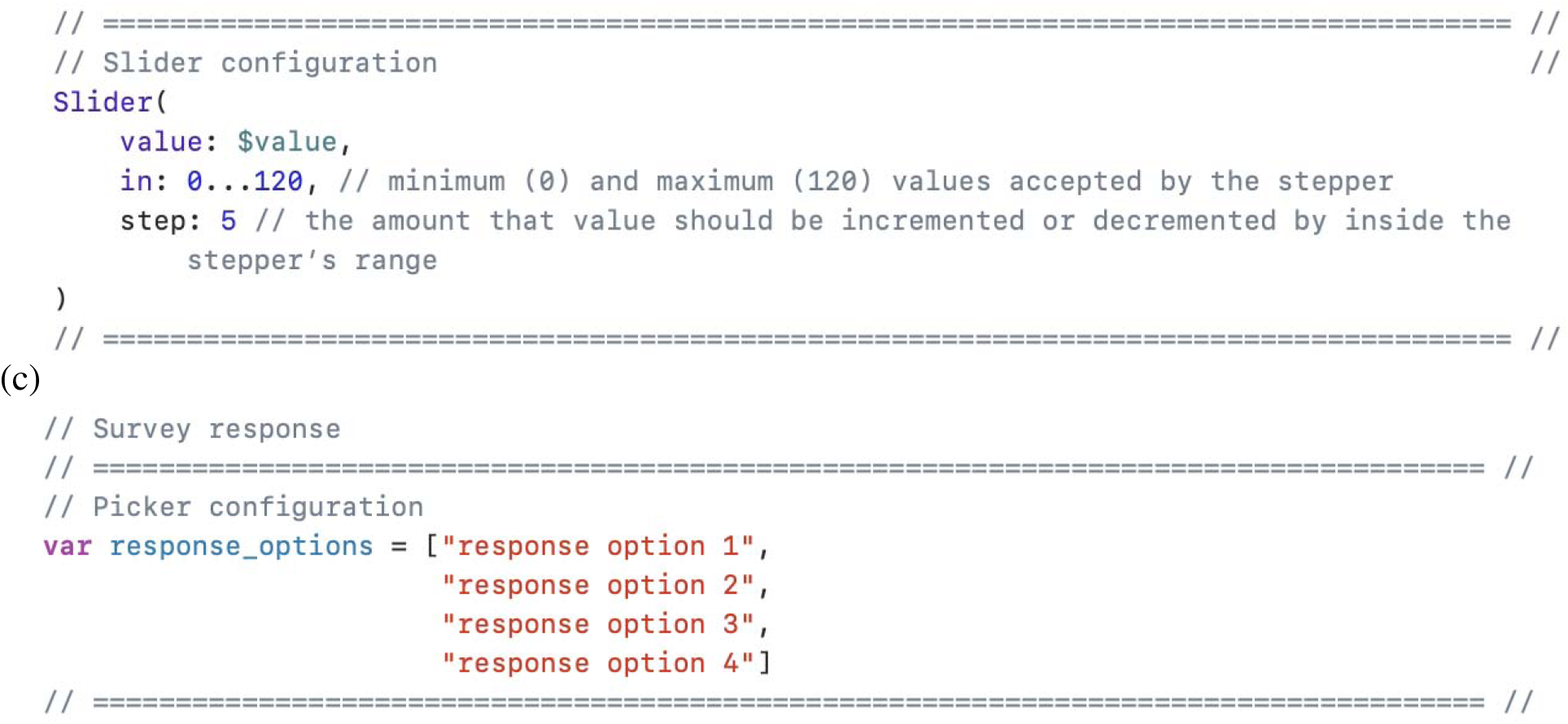
Survey question (a), slider configuration (b) and picker configuration (c).

#### 1.4 Views

##### Survey Selection

The Views directory encompasses the entire app interface and crucial mechanics of the app. Within this directory, there are various views available, and one of them is the `MainView`. The `MainView` includes a function called `getRandomSurveyView` (**Figure 5**) that allows for the selection of specific survey modules for future survey deliveries. By default, all five modules are included in the function, but it can be easily updated by keeping or removing lines that include the specific name of the desired survey module. The surveys are randomly chosen and delivered at random times within pre-defined time windows throughout the specified study period.

**Figure 5.**
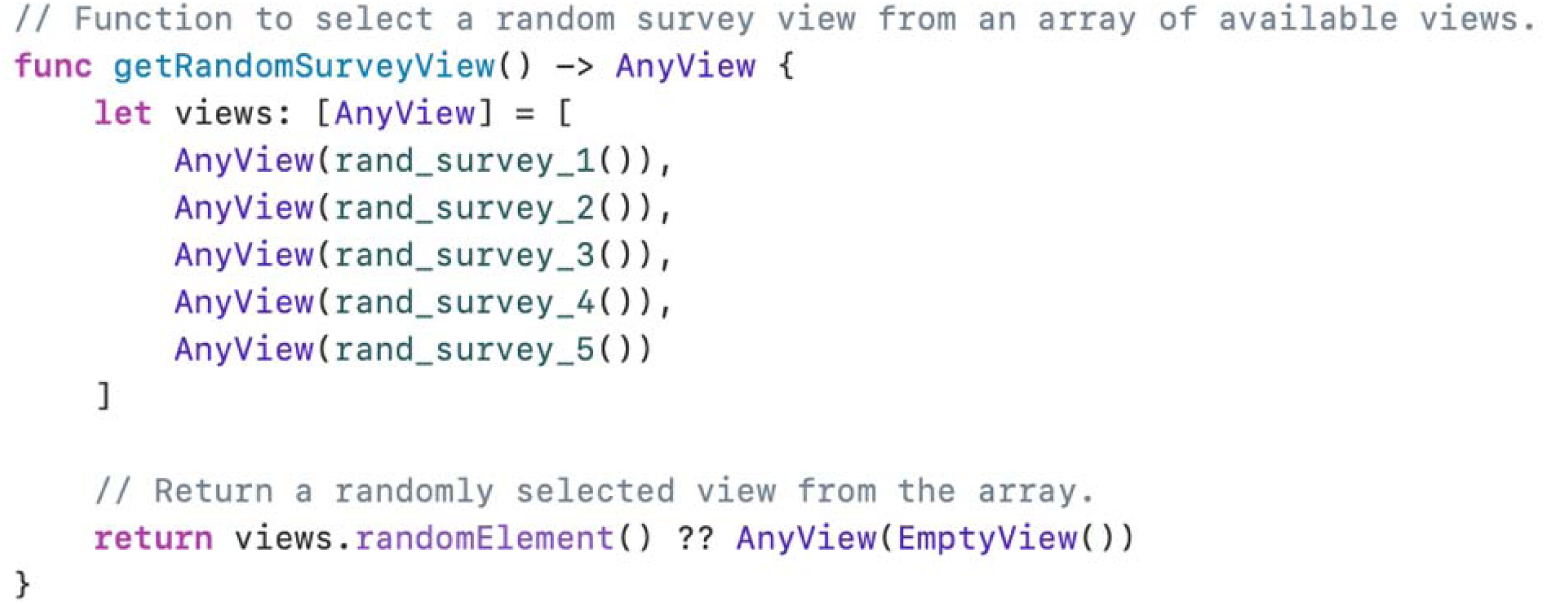
Survey selection.

##### Survey Schedule Display

The `MainView` also defines the layout of the app’s main screen. The DOSE framework app’s main screen comprises three primary elements: the User ID display, an upload button for uploading HealthKit data, and a calendar icon button for future survey notification schedule display (**Figure 6-a**). While hiding the survey delivery time is a standard practice in ESM studies to minimize bias and maintain ecological validity, including a survey delivery schedule display button in prototype ESM apps can be valuable for testing purposes. It facilitates debugging, customization, and experimentation with survey timing. However, once testing is complete, it is recommended to remove the display button from the actual ESM survey app before deploying it. To remove the display button, we can use a block comment method. Typically this involves surrounding the relevant code section with block comment symbols, such as `/*` and `*/`, effectively commenting out the code. This way, the code will be temporarily disabled while preserving a backup of the original code. The responsible code chunk is located under the commented line “`// List of scheduled notifications (today and tomorrow`)” (**Figure 6-b**).

**Figure 6.**
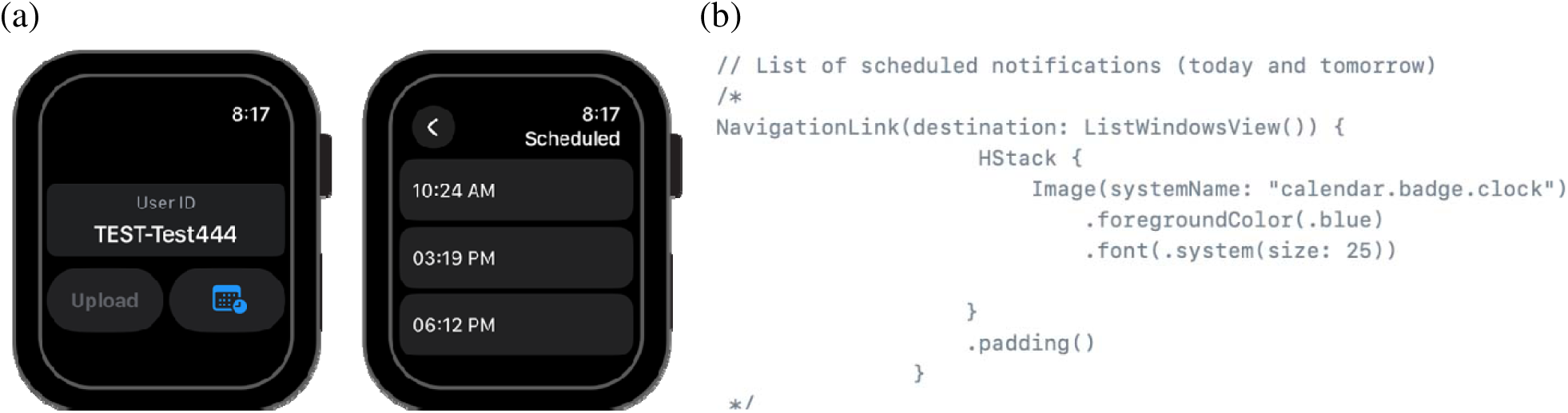
Survey schedule display on the app (a) and block-commented code for schedule display in the MainView (b).

##### App Deployment

Finally, by using the “Product – Archive” feature in Xcode, a new app for a native, ESM watch app can be created from the source code.

### Step 2. Participant Onboarding and Data Collection

#### 2.1. Push Notification Permission

After downloading the app from App Store or TestFlight, study participants are immediately prompted to grant notification permission (**Figure 7-a**). This permission is necessary as the app relies on push notifications to deliver EMS surveys to the participants.

**Figure 7.**
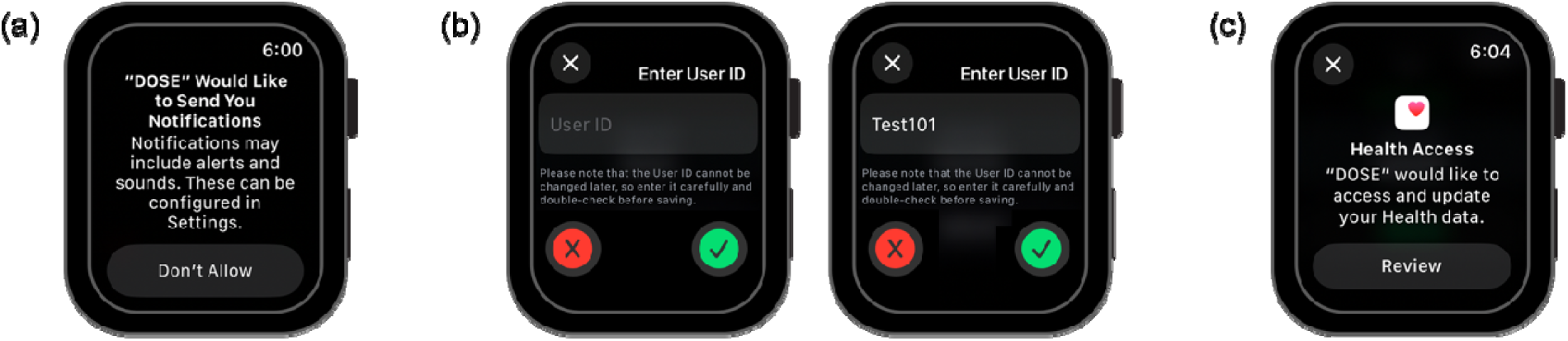
Notification permission grant (a), user ID selection (b), and HealthKit data access authorization (c).

#### 2.2 User ID Selection

Once notification permission is granted, study participants proceed to register with a User ID by typing it into a designated input box displayed on the top of the app’s home screen (**Figure 7-b**). The User ID can either be chosen by the participant themselves or assigned by the researchers. It must consist of both alphabetical letters and numeric values. This User ID is used to encrypt the data collected through the app. When first chosen, the User ID is linked to a unique identifier on each watch device, enabling device-specific tracking for HealthKit data collection throughout the study. Therefore, once selected, the User ID cannot be changed until the study period for that participant ends, unless the participant uses a new device. All data are linked to the chosen User ID and backed up in the Firebase Database. If a study provides devices to participants and redistributes a device to another participant after initial use, the device must be reset before a new User ID can be assigned.

#### 2.2. Authorizing Access to HealthKit Data

HealthKit serves as a central repository for health and fitness data on iOS. Apple provides an API that enables developers to request various types of continuous data, including step counts, heart rates, calories burned, and sleep patterns. Just like notification permission, when integrating HealthKit capability into an app, the app needs to request permission from the user to access and retrieve data from HealthKit, as well as to transfer it to a database. These permissions can be obtained by tapping the “`Upload`” button located at the bottom left of the app’s home screen (**Figure 7-c)**. Once the permissions are granted, the app can periodically retrieve data from HealthKit. In this framework, specifically, the step count and heart rate data are retrieved and transferred to Firebase Realtime Database with the User ID whenever the “`Upload`” button is tapped. Upon tapping, the data collected between the previous tap (or the last upload point) and the current tap (the present time) is fetched and transferred. This retrieval must be initiated manually by the user by pressing the “`Upload`” button. While automatic background retrieval, with a researcher-defined period, is planned for future versions, it has not been implemented in the current version of the framework.

### Step 3. Data Processing

#### 3.1. Firebase Directory Structure

With the DOSE framework app, all survey responses and HealthKit data are transmitted to Google Firebase Realtime Database in real-time as users submit or upload them. In Firebase Realtime Database, data is organized in a tree-like structure of nodes and keys. A node acts like a folder and can contain values or other nested nodes, while a key is a label identifying each piece of data. In the DOSE app, all data is organized under a single parent node called “`devices`,” which contains child nodes named after each participants’ User ID. Each of these child nodes has three additional child nodes: “`healthkitdata`”, “`notifications`” and “`responses`” (**Figure 8**). These nodes are automatically created when a study participant opens their survey notifications or upload HealthKit data for the first time.

**Figure 8.**
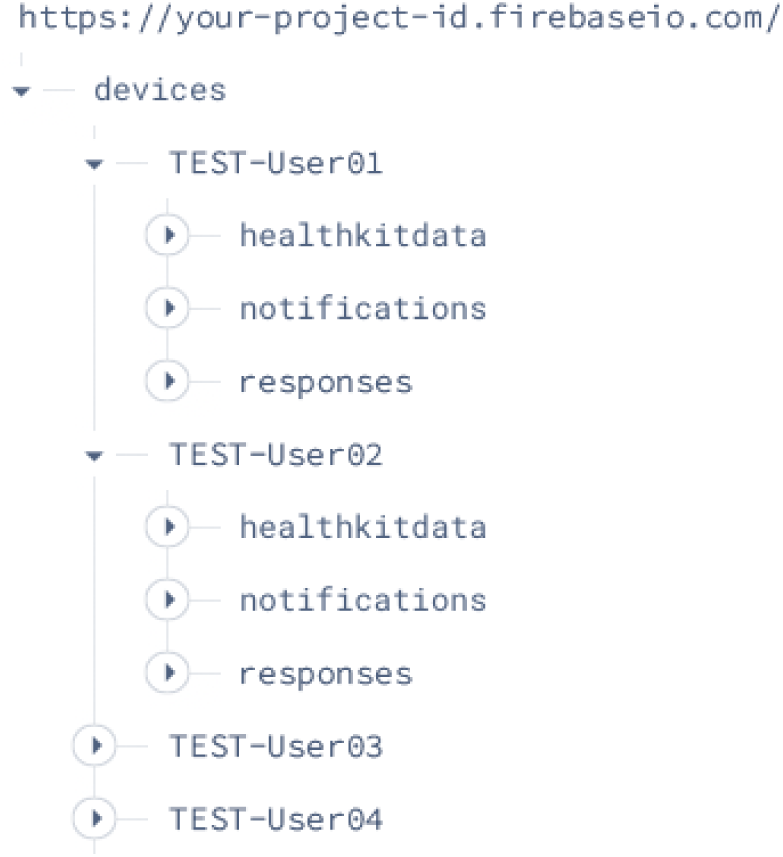
Firebase directory structure with parent and child nodes

#### 3.2. Survey Response and Notification Data

The response data for the ESM survey is stored in two nodes in Firebase Realtime Database: “`responses`” and “`notifications`” (**Figure 9**). The “`responses`” node contains keys for surveys that have been submitted or dismissed, including date and time information for when the survey was delivered, opened, submitted (or dismissed), along with the response value and survey module number. If a previous survey has not been completed and a new survey is delivered, the previous survey is marked as “expired” when the new survey is sent. Even if a response is later submitted for the previous survey in this scenario, it remains marked as “`expired`” in the survey module number field. The “`notifications`” node stores keys for surveys that were delivered and opened but never submitted or dismissed. It contains the date and time information for when the notification was delivered and opened. The purpose of the “notifications” node is to facilitate additional investigation into participants’ non-compliance behaviors.

**Figure 9.**
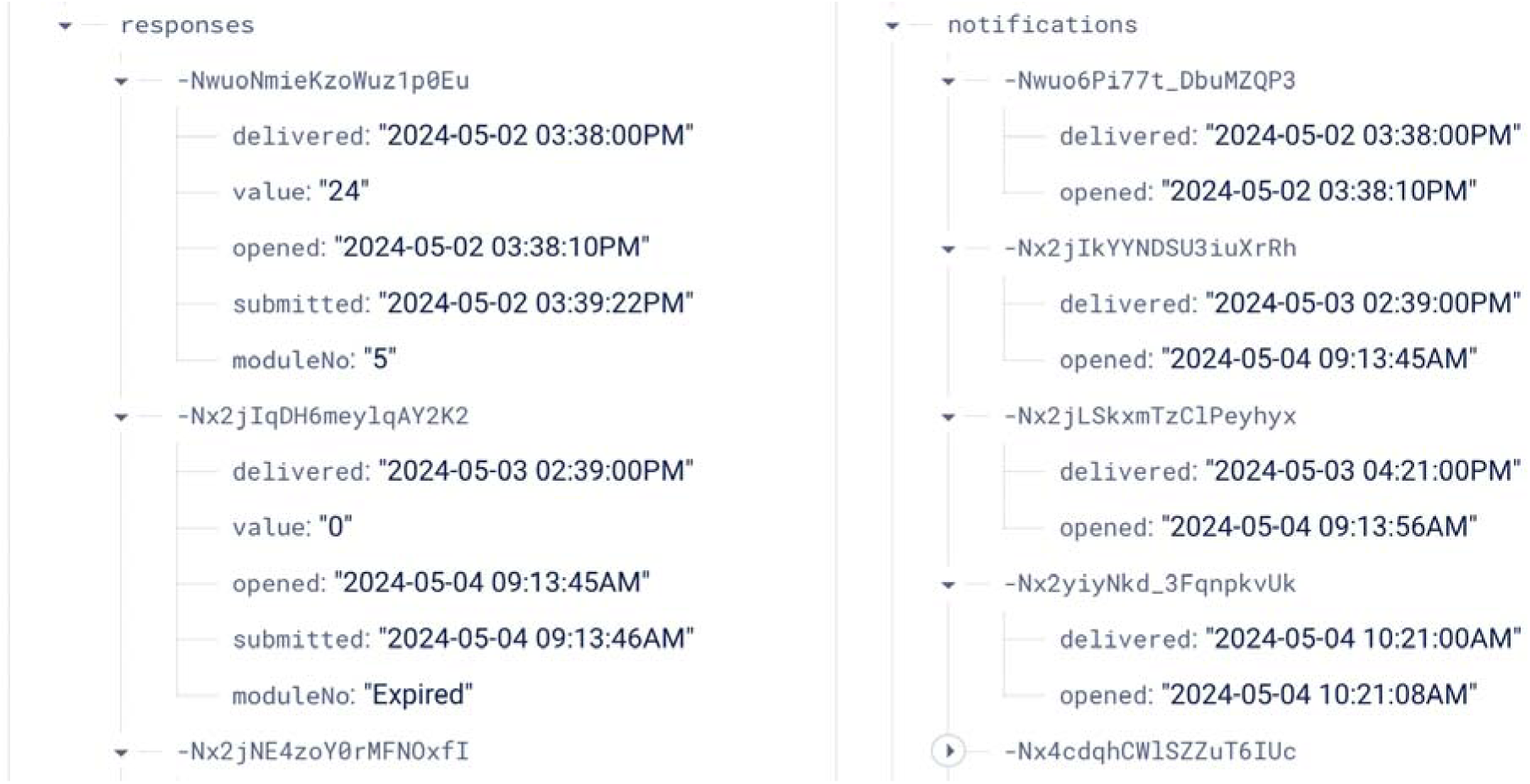
Survey response and notification data structure in Firebase with parent nodes, child nodes, and keys.

As of June 2025, the Apple Watch does not provide a direct way to track or retrieve notification delivery status or timestamps for notifications without user interactions, such as opening the notification. For instance, if a user swipes the notification to remove it, it is not possible to track the notification delivery status. The DOSE framework relies on the `UNUserNotifications API` to locally schedule notifications. Because notifications are stored directly on the device once scheduled, their delivery does not depend on network connectivity, which is the primary known cause of notification loss (An et al., 2016). Under normal conditions, this approach ensures virtually 100% reliability—a locally scheduled notification might not appear only if user has disabled notifications for the DOSE app or if the device is in a Focus mode (such as do not disturb, sleep, or work) (Apple Inc., n.d.-b).

#### 3.3. HealthKit Data

HealthKit records data as events of variable durations with discrete start and end times. The “`healthkitdata`” node consists of four keys: “`startDateTime`,” “`endDateTime`,” “quantity,” and “type.” Within these files, step counts and heart rates from HealthKit data are stored as raw, non-aggregated information (**Figure 10**). They are represented using predefined structures that include start date/time, end date/time, type (e.g., step counts, heart rates) and quantity. In HealthKit, the time intervals at which raw step counts and heart rates are measured can vary depending on the device. Different devices have varying capabilities and settings for capturing these data, with intervals ranging from a few seconds to a longer duration in 10-15 minutes. The `startDateTime` and `endDateTime` record the interval within which a participant’s device captures the data. The unit of measurement associated with the quantity is based on the participant’s device settings, allowing for potential variations in the units associated with HealthKit data according to user preferences. The quantity within these data structures represents the measure value between the `startDateTime` and `endDateTime` (e.g., 100 steps [2024/02/03 11:23 AM – 2024/02/03 11:34 PM], 1.6 beats per second [2024/02/03 1:41 PM – 2024/02/03 1:44 PM).

**Figure 10.**
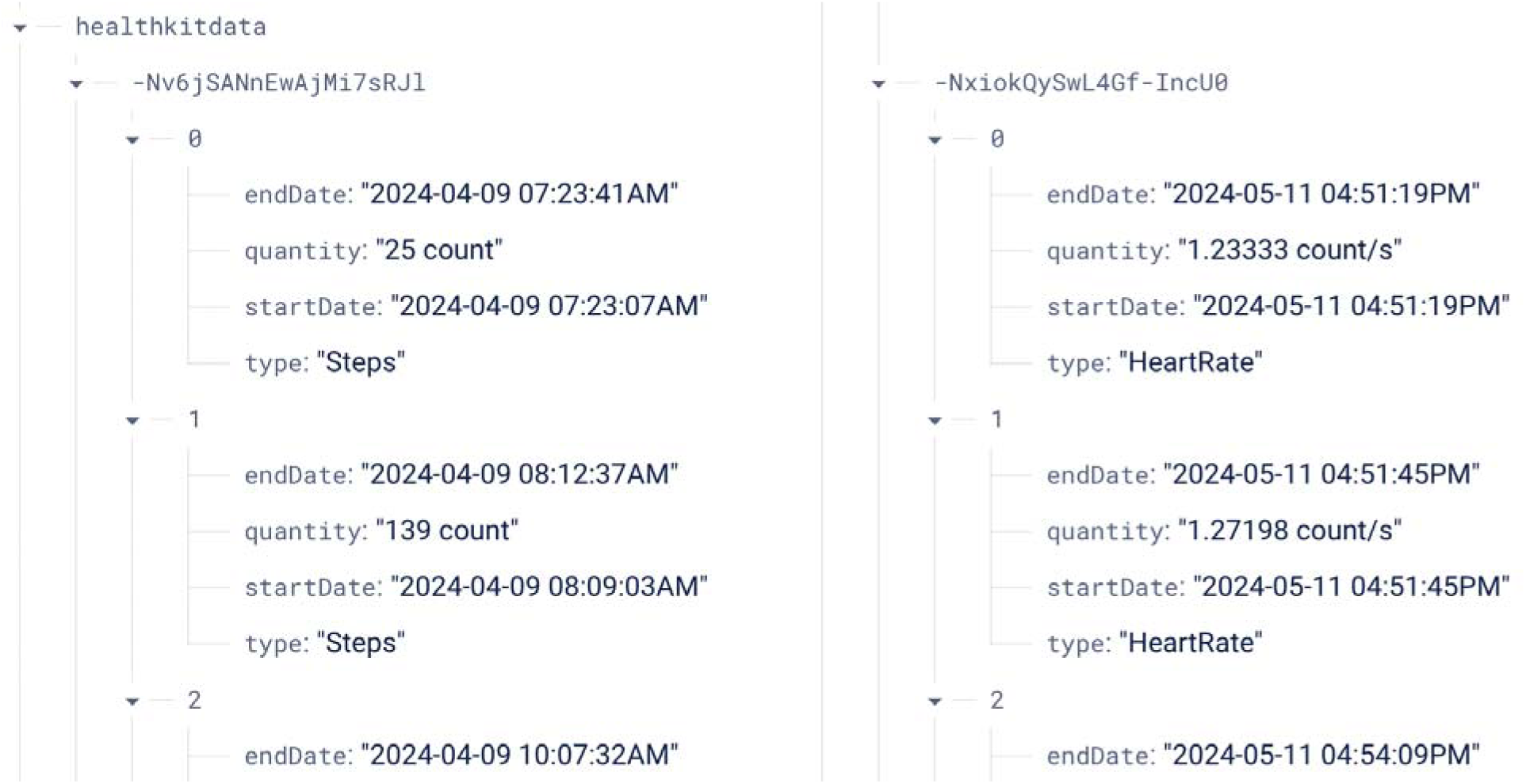
HealthKit data structure in Firebase.

#### 3.4. File Conversion From JSON to CSV

In Firebase Realtime Database, data is organized in a hierarchical tree structure, and each node within the tree can store data in JSON format. To download the JSON data from the desired location in the database tree, you can access the Firebase Realtime Database console and click on the “Export JSON” button, typically found at the top-right corner of the data view panel. This initiates the download of a file called “database-export.json” containing the selected data. It is worth noting that while the JSON format allows for flexible organization and representation of data within the console, it may not be optimal for certain types of analyses. To address this, we have developed a Python script specifically for researchers to download, process, and convert the data to a flat CSV file for analysis. The script, along with detailed instructions, can be downloaded via GitHub (https://github.com/iansulin/umich_dose).

## FEASIBILITY STUDY

### Objectives

We conducted a feasibility study to determine if it is possible to obtain valid data from Apple Watch-based ESM data collection using the new DOSE app. This app delivered random survey notifications (3 times daily) over a 28-day experience sampling period (84 prompts total) that exceeded the current iOS limit on scheduling notifications. Secondary objectives included assessing overall survey completion rates and evaluating device access time and interface usage time during micro-interactions. Micro-interactions are typically understood to be surveys that can be completed within 4 seconds or less (Intille et al., 2016; Ponnada et al., 2022). Access time refers to the duration from when a survey prompt is delivered to when the survey is opened, while interface usage time refers to the duration from when the survey is opened to when it is submitted. The study was approved by the Pennsylvania State University IRB (STUDY00023288).

### Recruitment and Participants

We recruited 30 adults (aged 40-80 years) who owned an iPhone and an Apple Watch with iOS v16 (or higher) and watchOS version 8.1 (or higher) through online social media platforms between January and October 2024. Participants were excluded if they reported 90 minutes or more of moderate-vigorous intensity physical activity in the past week, any contraindications to physical activity on the Physical Activity Readiness Questionnaire for Everyone (PAR-Q+; Warburton et al., 2011), had any mobility restrictions that interfered with unassisted physical activity, were pregnant or planning to become pregnant during the study, were concurrently participating in another study involving physical activity or weight loss, planned to have surgery or relocate during the study, or had been diagnosed with mild cognitive impairment or Alzheimer’s disease or dementia. Participants were randomly assigned with a 2:1 allocation ratio to use the DOSE app or not and this study uses data from the participants who used the DOSE app. Two participants from the DOSE app group withdrew within 24 hours without providing a specific reason. As a result, data from the remaining 18 participants were used in the analysis. Participants were compensated up to $250 for completing assessments (including activity monitoring and cognitive function assessments not reported here) but none of the payments were contingent on using or responding to prompts from the DOSE app.

### Survey Questions and Scheduling

For experience sampling, two survey questions were created: (1) “In the past 2 hours, how many minutes did you spend *exercising*?” and (2) “In the past 2 hours, how many minutes did you spend *moving*?” Each of these questions was paired with the five survey interfaces introduced in the 1.3 SurveyModule section, resulting in a total of ten surveys in the SurveyModule. The scrollable list interface offered options ranging from 0 to 120 minutes in 5-minute intervals. The step slider interface allowed users to increase or decrease values within the same range and intervals using + or – buttons. The circular slider interface also provided options from 0 to 120 minutes, with increments or decrements of 1 minute. All interfaces defaulted to 0. During the 28-day study period, notification prompts were delivered three times per day over a 12-hour period, between 8 AM and 8 PM. These prompts were randomly distributed across three 4-hour windows throughout the day. Among the ten possible survey configurations (= 2 questions X 5 interfaces), only one was randomly selected and delivered at a time. Notifications were scheduled for ten days, with three notifications each day, totaling 30 notifications at once. Every 3 days, a new set of 30 notifications was automatically rescheduled.

### Measures

Survey completion rates for the experience sampling were calculated as a proportion of the number of surveys submitted out of the number of surveys scheduled. To capture behaviors in real-world settings, participants were not instructed to keep their devices unusually charged or wear them more frequently than usual; instead, they were asked to maintain their regular Apple Watch usage. As a result, some participants occasionally forgot to charge their watches or wear them on certain days. Consequently, days when no surveys were opened were classified as ‘non-wear days’ and excluded from the analysis. Micro-interaction rates were calculated as the percentage of interface usage lasting 4 seconds or less.

### Results

The study app, built on the DOSE framework, successfully delivered randomized survey prompts three times daily within predefined 4-hour time windows over the 28-day study period, totaling 84 notifications—surpassing the standard iOS scheduling limit of 64 (**Figure 11**). Surveys were delivered without technical interruptions or scheduling failures, demonstrating both the feasibility of our extended notification scheduling approach and the practicality of using DOSE framework for experience sampling.

**Figure 11.**
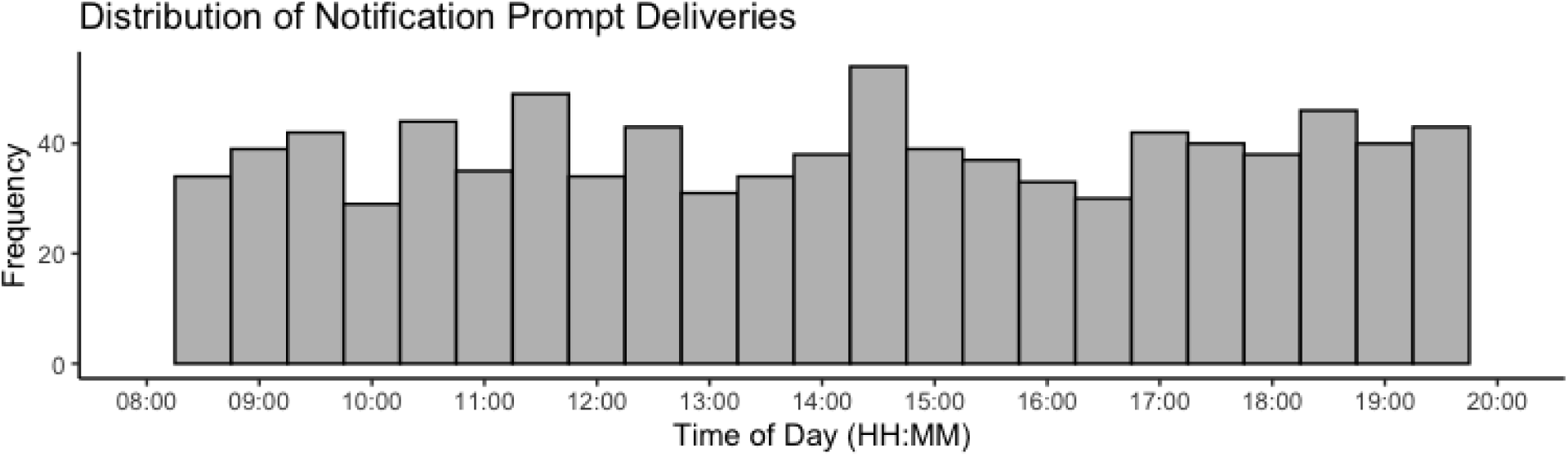
Distribution of notification prompt deliveries

On valid days—when participants wore their Apple Watch—survey completion rates averaged 76% for middle-aged adults and 86% for older adults (**Table 2**). Despite higher completion rates, older adults had fewer valid days than middle-aged adults (but this finding appeared to be driven by three participants who struggled with watch wear). Among incomplete surveys, the proportions that were opened but not submitted—possibly due to interruptions, technical difficulties, or accidental dismissal (e.g., clumsy hands)—was 16% for middle-aged adults and 27% for older adults. Most middle-aged participants showed relatively consistent behavior, with occasional missed surveys or device non-wear; older adults, however, showed more variability: some rarely wore the watch, while others wore it consistently and completed most surveys (**Figure 12**).

**Figure 12.**
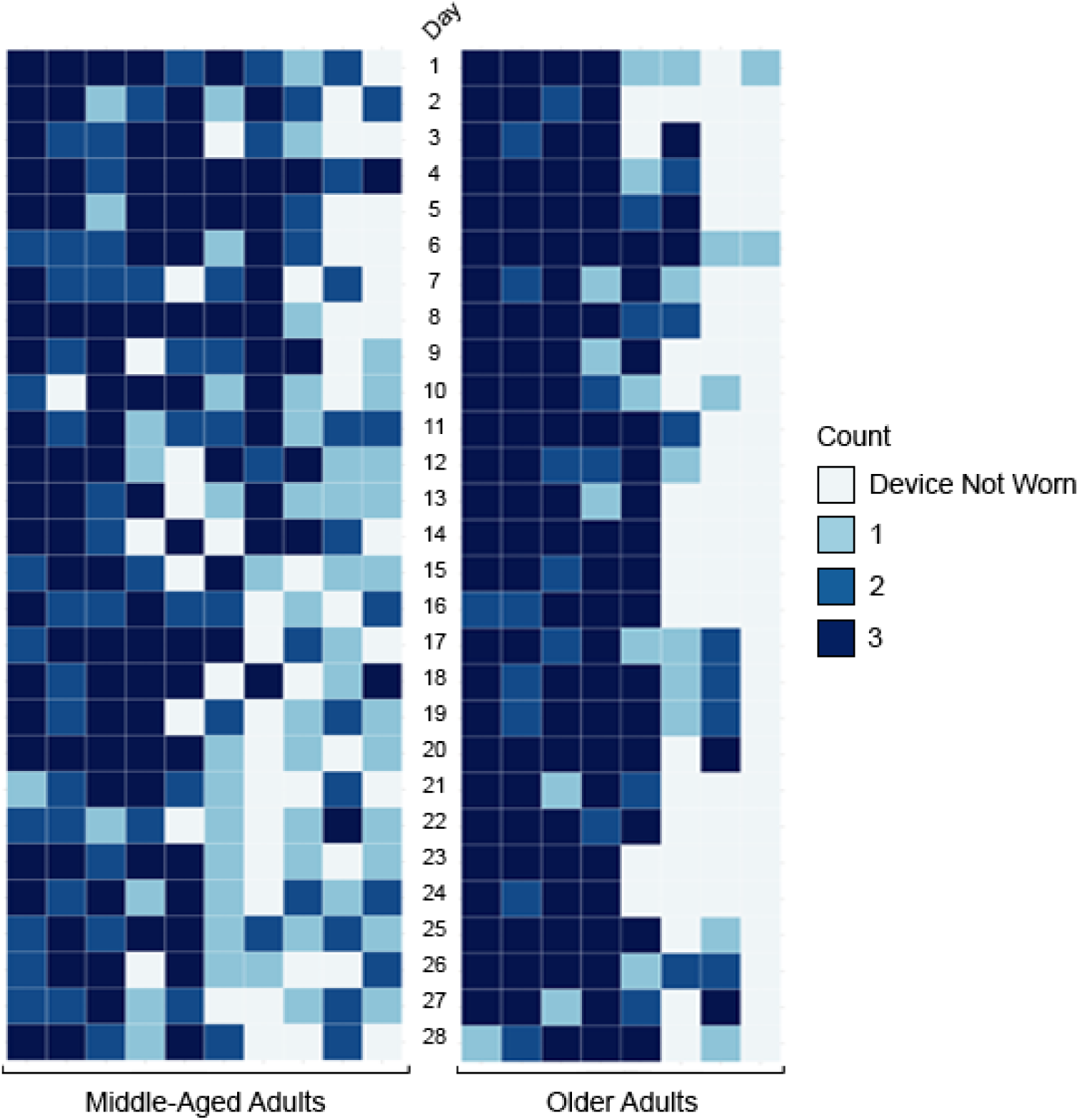
Heatmaps showing the number of daily completed surveys by participants over 28 study days. The vertical axis represents study days (1 to 28), while each column corresponds to an individual participant. Color intensity indicates the survey completion frequency, with darker shades representing higher counts.

**Table 1.**
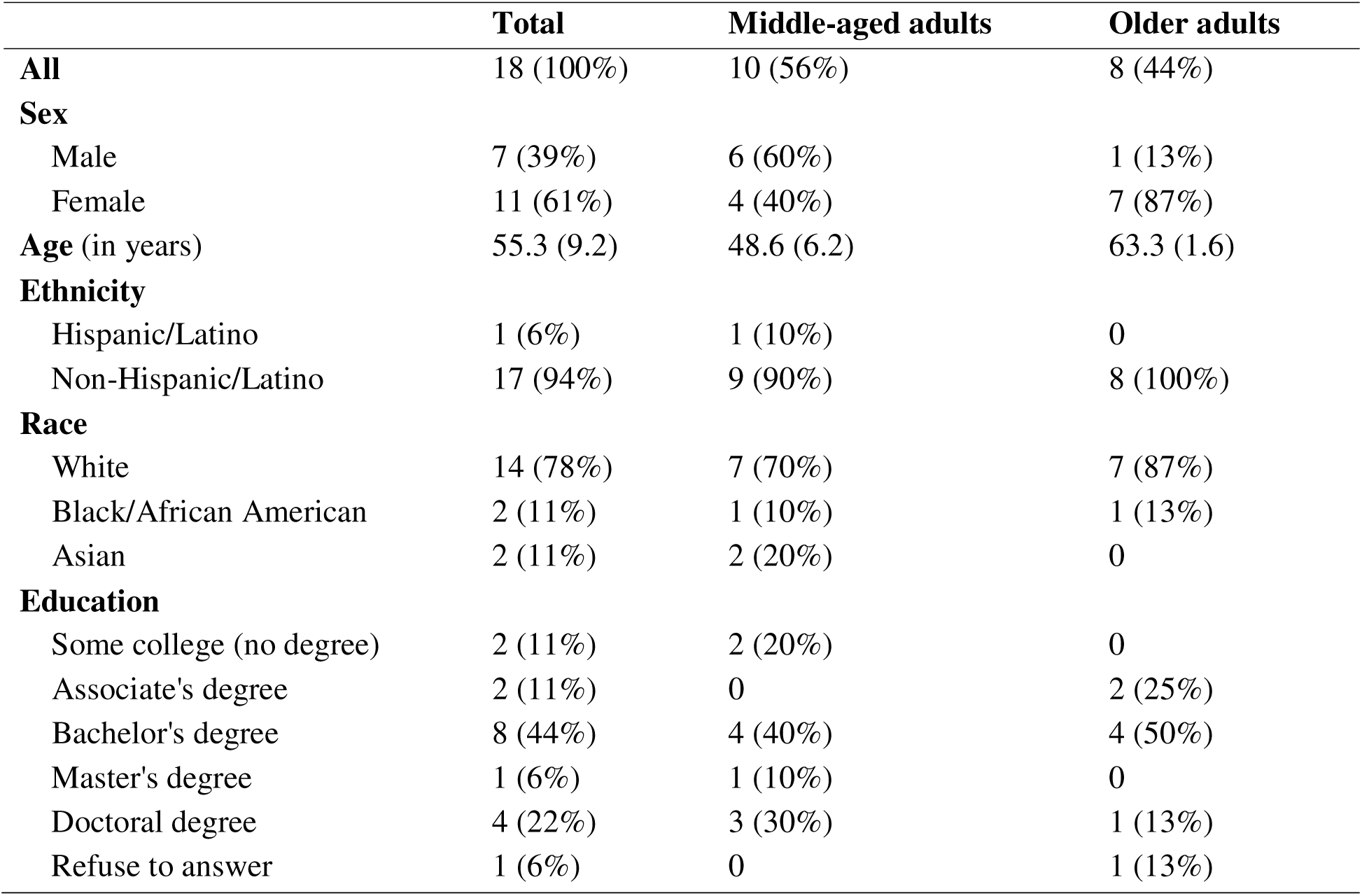
Demographic characteristics of sample that completed the study (N = 18)

|  | **Total** | **Middle-aged adults** | **Older adults** |
| --- | --- | --- | --- |
| **All** | 18 (100%) | 10 (56%) | 8 (44%) |
| **Sex** |  |  |  |
| Male | 7 (39%) | 6 (60%) | 1 (13%) |
| Female | 11 (61%) | 4 (40%) | 7 (87%) |
| **Age (in years)** | 55.3 (9.2) | 48.6 (6.2) | 63.3 (1.6) |
| **Ethnicity** |  |  |  |
| Hispanic/Latino | 1 (6%) | 1 (10%) | 0 |
| Non-Hispanic/Latino | 17 (94%) | 9 (90%) | 8 (100%) |
| **Race** |  |  |  |
| White | 14 (78%) | 7 (70%) | 7 (87%) |
| Black/African American | 2 (11%) | 1 (10%) | 1 (13%) |
| Asian | 2 (11%) | 2 (20%) | 0 |
| **Education** |  |  |  |
| Some college (no degree) | 2 (11%) | 2 (20%) | 0 |
| Associate's degree | 2 (11%) | 0 | 2 (25%) |
| Bachelor's degree | 8 (44%) | 4 (40%) | 4 (50%) |
| Master's degree | 1 (6%) | 1 (10%) | 0 |
| Doctoral degree | 4 (22%) | 3 (30%) | 1 (13%) |
| Refuse to answer | 1 (6%) | 0 | 1 (13%) |

**Table 2.** Overall survey completion rates over the 28-day study period and number of valid days by age group.

| Group | Overall Survey Completion Rate (%) |  | Number of Valid Days (Watch Device Worn) |  |
| --- | --- | --- | --- | --- |
|  | Mean (SD) | Median (IQR) | Mean (SD) | Median (IQR) |
| Middle-Aged Adults | 75.59 (9.54) | 75.54 (10.81) | 22.90 (4.20) | 23 (7.50) |
| Older Adults | 86.02 (9.02) | 93.33 (12.62) | 20.13 (10.32) | 26 (15.75) |

**Figure 13** shows the weekly completion rates, and distributions of time spent on device access, interface usage, and total response by age group. Over the four-week period, completion rates among middle-aged adults declined slightly, while rates for older adults remained relatively stable. The median device access time was 9 seconds (IQR = 61) for middle-aged adults and 9.5 seconds (IQR = 1241) for older adults. The median interface usage time was 10 seconds (IQR = 14), with a micro-interaction rate of 24.1% (SD = 19.0%) for middle-aged adults, and 13 seconds (IQR = 11), with a micro-interaction rate of 6.9% (SD = 6.5%) for older adults. The median total response time was 22 seconds (IQR = 73.5) for middle-aged adults and 29 seconds (IQR = 1256) for older adults. For both middle-aged and older adults, all three time metrics—device access time, interface usage time, and total response time—consistently decreased across the weeks, suggesting that participants became more efficient with the technology and survey procedures through repeated use. Although we did not statistically compare these weekly trends between groups, due to limited power from the small sample size, visual inspection suggests that middle-aged adults appeared to adapt more quickly than older adults.

**Figure 13.**
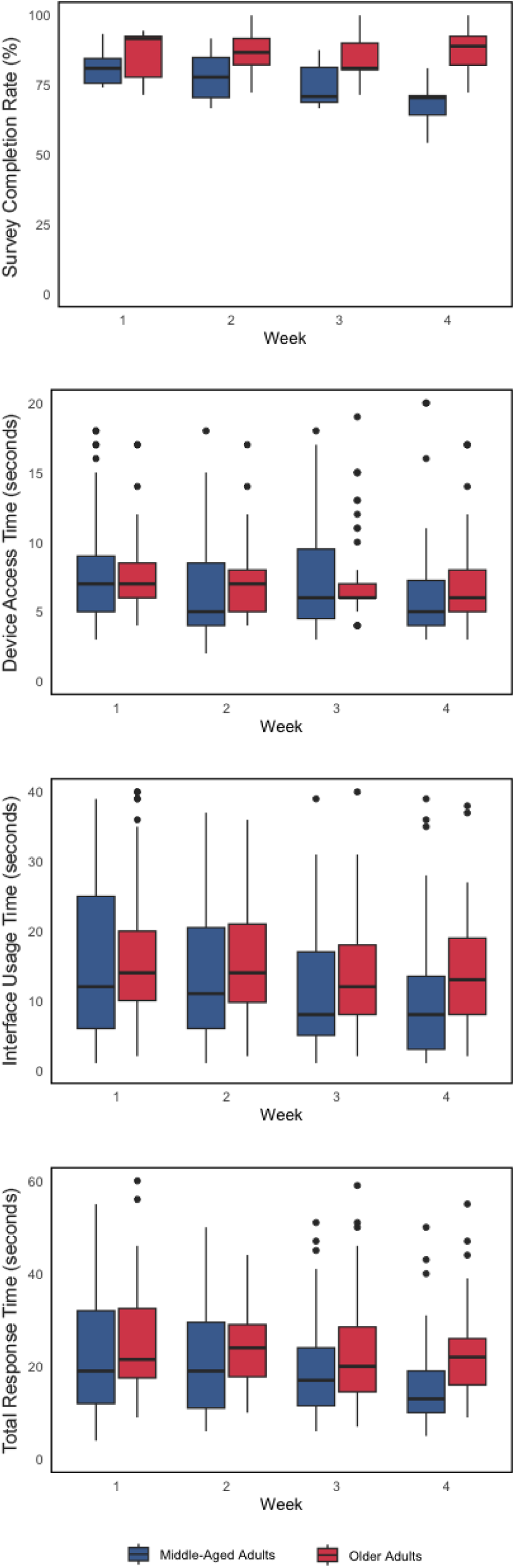
Box plots showing weekly completion rates by age group.

## DISCUSSION

This paper introduced DOSE, a new app for research using ESM on Apple Watches and reported on the feasibility of deploying the app with middle-aged and older adults. The wearable nature of smartwatches makes them an intriguing method platform for ESM but idiosyncrasies in the Watch iOS have limited applications of the Apple Watch for experience sampling schedules with more than 64 randomly-scheduled prompts. The new DOSE app for Watch OS circumvents that limitation and enables researchers to deliver randomly-scheduled prompts without an upper limit on study duration. Feasibility testing revealed survey completion rates above 75% for both middle-aged and older adults. Older adults appeared to not wear their watches on some days so device wear will be an important consideration when developing sampling protocols for older adults. Responses were relatively quick following prompts for both middle-aged and older adults; however, older adults had some prolonged response times so future deployments should verifying that users activate notification permissions from their Watch. Participant generally completed responses within 30 seconds of receiving a prompt. Over the four-week test period, both middle-aged and older adults appeared to become more proficient with the app as evidenced by faster response times. This work makes three significant contributions to the literature.

First, the DOSE app facilitates broader implementation of ESM via smartwatches by successfully circumventing a limitation of the Apple Watch OS that precluded extended sampling schemes. Smartphones are currently the dominant technology for conducting research with ESM. The first smartwatches were introduced to consumers just over a decade ago and have since emerged as an industry with over $10B sales in the US and over $35B (Fortune Business Insights, 2023). Smartwatches offer one key advantage over smartphones – they are designed to be worn on the person which increases proximity for responding to notifications. One barrier to using smartwatches for ESM data collection is the limited number of suitable apps. For Android devices, researchers have developed several options tailored to their needs (Intille et al., 2016; Kim et al., 2022; Ponnada et al., 2021). The fragmented nature of the Android smartwatch ecosystem—where apps developed for one device are often incompatible with others—have often led researchers to focus on one or a few specific Android smartwatch models or non-commercial devices (Delobelle et al., 2025; Intille et al., 2016; Kim et al., 2022; Ponnada et al., 2022; Volsa et al., 2022). For Fitbit smartwatches, Fitabase developed a platform called Engage to enable ecological momentary assessments (including ESM) (Conroy et al., 2023); however, we are not aware of any open-source solutions for those devices. The DOSE app provides a new and, to the best of our knowledge, the first, open-source option for conducting ESM research on the Apple Watch over long time intervals. The DOSE framework specifically addresses these challenges through custom functions for dynamic notification scheduling and random date/time selection, enabling researchers to configure study duration, time windows for notifications, and prompt frequency via user-defined parameters tailored to specific research needs. As the first open-source framework for developing ESM applications on Apple Watch, DOSE not only expands platform accessibility but also enhances the inclusivity, flexibility, and long-term applicability of ESM research.

Prior research on smartwatch use for experience sampling has largely focused on young and middle-aged adults. Developing technologies for older adults will be essential for facilitating their representation in research. A major demographic transition is underway in the US and the Census Bureau projects that the country will have more older adults than children or youth within a decade (U.S. Census Bureau, 2018). Despite many negative stereotypes about aging adults’ technology use, mobile technology adoption continues to increase among middle-aged and older adults (AARP, 2025; Pew Research Center, 2022). Adoption of wearable technologies, such as smartwatches, lags behind somewhat but will inevitably catch up as existing older adults adopt new technologies and younger cohorts who have adopted those technologies age in (Farivar et al., 2020; Lazaro et al., 2020; Manini et al., 2019).

Results from our feasibility study add to accumulating evidence that older adults can use these devices. For example, one recent study explored the potential of smartwatches for ESM in older adults, finding that participants generally viewed smartwatch technology positively and expressed interest in its real-time assessment capabilities, but concerns about small screens and interface complexity were noted (Manini et al., 2019). Similarly, another study reported that while general perceptions of seniors’ use of smartwatches often focus on cognitive aging, older adults themselves identified the difficulty of reading and interpreting smartwatch displays as a key barrier to adoption (Farivar et al., 2020). While two recent studies explored the use of smartwatches with older adults, the devices were primarily used as activity trackers in lab-based setups, with no experience sampling conducted (Delobelle et al., 2025; Holmqvist et al., 2025). To our knowledge, our study is among the first to employ smartwatches for experience sampling in middle-aged and older adults, establishing the real-world feasibility of smartwatch-based ESM studies in this population. Our study demonstrated that middle-aged and older adults are both willing and able to engage with an Apple Watch app for self-monitoring. The greatest challenge for future work with older adults involves promoting daily device wear; however, we caution that these results were based on a small sample and could reflect idiosyncrasies of the eight older adults in the study rather than a generalizable characteristic of older adults. Slight increases in response times compared to prior work likely resulted from the naturalistic design of our study (Intille et al., 2016). Unlike prior research with strict constraints—such as 20-second response limits and standardized 3-second vibration prompts— participants used their Apple Watches under typical conditions, including personalized vibration settings and occasional use of silence mode. The DOSE app also featured five distinct survey interfaces with two different questions, which may have contributed to longer response times by encouraging more deliberate reflection and requiring prolonged interactions to navigate a broader range of response options within each interface—unlike prior studies that used a single interface with one question and only 2–3 visible options, allowing for quick, repetitive one-tap responses. These findings provide encouraging evidence for the practical, real-world applicability of smartwatch-based ESM in middle-aged and older adults, expanding the reach and inclusivity of future research. Overall, these findings provide further evidence to challenge ageist beliefs that have contributed to a digital health divide for older adults (Mace et al., 2022).

Our findings also underscore the importance of user behavior as a key factor influencing ESM compliance, particularly among older adults who appeared to face challenges in consistently wearing or charging their devices. Irregular charging routines can lead to missed prompts due to power loss, preventing scheduled notifications from triggering and resulting in expired surveys. Additionally, tactile notifications can be easily missed during physical or cognitively demanding activities. While these factors present challenges, they also offer opportunities for future research to develop more user-specific and context-aware prompting strategies. Nevertheless, when devices were worn and prompts were detected, participants in our study demonstrated consistently low response latency—9 seconds for middle-aged adults and 9.5 seconds for older adults—suggesting that smartwatches can support timely ESM engagement, even among populations that may face usability barriers. For comparison, smartphone-based ESM studies have reported median latencies exceeding 3 minutes (Conroy et al., 2020; Delobelle et al., 2025). This low latency likely reflects the reduced friction of smartwatch access, greater tactile prompt salience, and a more lightweight interaction design—all of which together contribute to near-instantaneous engagement. Future studies built on the DOSE framework may further explore and characterize individuals’ temporal engagement patterns with ESM in relation to device use behaviors and contextual factors, with the goal of optimizing prompt delivery and further enhancing compliance.

A third contribution of the DOSE app is that it can be extended to deliver some ecological momentary interventions that require prompting. One such intervention is self-monitoring, a well-established behavior change technique that involves observing and recording one’s behavior (Marques et al., 2024). This technique has a strong evidence base in the physical activity promotion literature (Samdal et al., 2017). Recent results have suggested that prompting users to reflect on their activity levels 3-4 times per day is associated with increased daily physical activity, whereas the frequency of passive behavioral feedback showed no such association (Conroy et al., 2023). Similarly, future research may also benefit from incorporating self-regulation prompts or learning opportunities to enhance users’ self-management capabilities. A recent review further supports this approach, indicating that brief, message-based prompting interventions can significantly improve both preventive health behaviors (e.g., smoking cessation) and clinical outcomes (e.g., diabetes management) (Fjeldsoe et al., 2009). Moving forward, investigations into the optimal timing, frequency, and content of such prompts may help maximize user engagement and intervention efficacy. As such, the DOSE framework offers a foundation for new tools for delivering ecological momentary interventions that promote active engagement in healthcare and evidence-based behavior change strategies.

The DOSE app has several limitations. Perhaps most obviously, this app was written to work around limitations of the Apple Watch OS. It is possible that Apple Watch users differ systematically from users of other devices (e.g., Fitbit, Garmin, Google, or Samsung). The code will also not work on other operating systems so it would need to be paired with a companion app for other devices used in a study. Researchers sometimes provide study devices for participants to use during a study, the Apple Watch is not a low-budget device so this solution may not be possible for all researchers. Second, the app does not track prompts that are ignored and not opened. In our testing, the app scheduled notifications reliably but ignored prompts cannot be distinguished from prompts when the user’s Watch is not worn. The app also does not currently allow different availability windows to be set for different days. Finally, the DOSE app was designed for ESM but some research questions require different sampling schemes (e.g., event-contingent recording). Likewise, although the app can deliver ecological momentary interventions on a random schedule, just-in-time adaptive interventions with tailoring variables and decision rules are not currently possible. The code would require more substantial modifications for other sampling schemes or to deliver just-in-time adaptive interventions.

In sum, the DOSE app is an open-source tool created to work around limitations for ESM research on the Apple Watch. This app provides another option for researchers interested in sampling momentary states in users’ lives. Whereas most prior work with smartwatches has focused on young adults, this study established the feasibility of obtaining high-quality data from middle-aged and older adults.

## Data Availability

The data collected for the feasibility study using the DOSE framework are not publicly available due to restrictions related to participant confidentiality. However, the data can be made available from the corresponding author upon reasonable request.

## DECLARATIONS

### Ethics approval

The study was approved by the Pennsylvania State University IRB (STUDY00023288).

### Consent to participate

All participants provided informed consent prior to participation in the study.

### Consent for publication

All authors have reviewed the final manuscript and consent to its publication.

### Availability of data and materials

The dataset generated during and/or analyzed during the current study are not publicly available due to restrictions related to participant confidentiality but available from the corresponding author on reasonable request.

### Code availability

The source code for the Stanford Screenomics platform developed by I.K. and S.K. is available for public access at: https://github.com/iansulin/umich_dose

### Authors’ contribution

I.K. conceived and designed the work, conducted data acquisition and analysis, interpreted data, drafted the manuscript, and performed critical revisions. I.K. and S.K. developed the software. S.A. contributed to the conception and design of the work. D.E.C. contributed to the conception and design, data interpretation, critical revisions, and secured funding. All authors reviewed and approved the final manuscript.

